# A Cross-Country Analysis of the Effectiveness of COVID-19 Vaccines in Reducing Mortality Rates within the EU

**DOI:** 10.1101/2022.01.23.22269604

**Authors:** Svetoslav Bliznashki

**Affiliations:** Sofia University, Sofia, Bulgaria

## Abstract

We use a linear mixed model in order to estimate the effect of the number of people vaccinated against COVID-19 on the overall death toll on a monthly basis. We limit our analysis for the duration of the year 2021 and within 25 countries which are current or former (UK) members of the EU since these countries follow similar approaches to testing and reporting different COVID-19 related statistics. We explored the effect in question by comparing the total number of people vaccinated up to the end of each month and the total number of deaths occurring during the next month while controlling for several measures including number of new COVID-19 cases, diabetes prevalence, cardio vascular death rates and time trends among others. Our results indicated that one percentage point monthly increase in the total number of vaccinated people was associated, on average, with a decrease of two deaths per general population of 1 million for the next month with the effect being highly significant. An Individual Growth Curves Analysis further corroborates these findings and suggests that vaccination rates may possibly exert additional indirect effects unaccounted for by our main model.

## 1. Introduction

The beginning of the year 2021 marked the beginning of the massive COVID-19 vaccination campaign within the EU. Currently, vaccines’ effectiveness with respect to disease prevention is coming into question (two possible reasons being anti-bodies waning over time and/or new variants exhibiting greater vaccine resistance); still, however, most analyses show that vaccines continue to provide significant protection with respect to serious complications, hospitalization and death (e.g. Pouwels et al., 2021; Hyams et al., 2021; Levin et al., 2021; see also COVID-19 Vaccine Surveillance Report Week 38 by Public Health England for a review). In order to investigate this last claim further, we chose 25 countries, current and former (UK) members of the EU with populations exceeding 1 million and analyzed the data for their cumulative vaccination trends and the effects of said trends on the deaths attributed to COVID-19 on a monthly basis.

Our inclusion criteria were based on the fact that EU countries tend to follow similar procedures and guidelines with respect to testing and reporting novel COVID-19 cases and mortality rates with their respective infrastructures presumably being capable of achieving relatively reliable estimates. The United Kingdom was also included in our analyses since its protocols and procedures don’t appear to diverge significantly from the ones employed in the EU, despite the country’s recent departure from the union. Countries with small populations (i.e. countries with less than 1 million people such as Luxembourg, Malta, etc.) were excluded in order to avoid biases with respect to population dynamics which may show aberrant trends in small and/or relatively isolated communities.

At least some analyses indicate that while in mild cases COVID-19 symptoms’ duration appears to be between one and two weeks, for severe and critical cases symptoms tend to last up to six weeks after their onset^1^. At the same time, most vaccines appeared to have measurable effects in reducing the risk of COVID-19 contraction 15 weeks after the first dose (e.g. Amit et al., 2021; Pilishvili et al., 2021) with the effects, presumably, being even larger with respect to complications and death. In that context, we judged it appropriate to conduct our analyses on a monthly basis, i.e. we tried to investigate the effect of the cumulative percentage of vaccinated people (i.e. people who had received at least one dose of any EU-approved vaccine) up to and including a given month on the total number of deaths due to COVID-19 for the next month (only) of the year^2^. In other words, we judged that a one month lag was a suitable interval for detecting the effect in question. For example, we recorded the total number of vaccinated people up to and including January 2021; then we recorded the total number of deaths due to COVID-19 which had occurred only during February 2021. The relationship between the predictor (cumulative percentage of vaccinated people) and the 1 month lagged outcome (the total number of deaths occurring only during the next month of 2021) is the primary focus of this study.

## 2. Materials and Methods

The data used in this study was retrieved from the *ourworldindata* repository^3^. We organized the data on a monthly basis. We organized the data from 25 European countries in a long format with each country encompassing 11 rows (i.e. we had 11.25=275 data points for our dependent measure across the entire dataset). The *cumulative percentage of vaccinated people* (i.e. this variable included only monotonically increasing values) up to and including the last day of each month (from January to November) was our main predictor (first column). Our second (control) predictor *was the total number of new cases of COVID-19* (per 10 000) which had occurred during a given month only (up to and including November). We also included a *linear* and a *quadratic time trend* in order to model basic and seasonal time variations in our outcome measure. The linear time variable (column 3) coded each successive month with numbers ranging from 0 to 10 (February=0, March=1, etc.); the quadratic time trend (time2) included the same numbers squared (i.e. it included eleven values ranging from 0 to 100) and was placed in the 4^th^ column^4^. Our dependent measure (column 5) was *the total number of deaths* (per million) *occurring during a particular month only*. The cumulative number of vaccinated people as well as the total number of new cases per month were recorded from January to November, while the time covariates and the dependent measure (total number of deaths per Million for a given month only) were recorded from February to December. In that way we had a dataset which comprised the whole 2021 year with a one month lag between our two main predictors (cumulative percentage of vaccinated people and number of new cases for a given month) and our outcome measure (total number of deaths due to COVID-19 during a given month).

In addition to our time-varying covariates described above, we also recorded several fixed covariates (i.e. variables which didn’t change in time within countries) which served as additional (fixed) controls. This data was obtained for 2019 and was comprised of the following variables: *population density, percentage of people over the age of 65, gross domestic product per capita, cardio-vascular death rate, diabetes prevalence, hospital beds per 1000 people*, and *life expectancy*.

The 25 European countries included in the analysis (in alphabetical order) were: Austria, Belgium, Bulgaria, Croatia, Czech Republic, Denmark, Estonia, Finland, France, Germany, Greece, Hungary, Ireland, Italy, Latvia, Lithuania, Netherlands, Poland, Portugal, Romania, Slovakia, Slovenia, Spain, Sweden, and the UK.

The vaccination rates for these countries varied between 0.41% (Bulgaria) and 13.63% (UK) for January and between 27.61% (Bulgaria) and 90.5% (Portugal) for December. In other words, it is clearly the case that 2021 provides us with enough variance in terms of vaccination rates both temporally and geographically in order to justify the analyses that follow.

Several linear mixed effects models were fitted to the data by the lme4 package for the open source R environment (Bates et al., 2015).

## 3. Analyses and Results

A linear mixed effects model (LMEM) fits a random intercept and random slopes for each unit of measurement for the different predictors which vary across the different measurement units. The random intercept/slopes suggest particular variance-covariance structures across different measurements within a unit (e.g. Singer & Willett, 2003).

In order to specify an appropriate covariance structure for our dependent measure we began by fitting a baseline model which included all our time-varying covariates – cumulative percentage of vaccinated people (%vac), number of cases per 10 000 (#cases), time and time squared (time2), as well as an interaction term for %vac and #cases (all covariates involved in interaction terms were centered^5^). The baseline model also specified a random intercept and random slopes for %vac and #cases within an unstructured covariance matrix (i.e. the variances of all random effects as well as the covariances between them were freely estimated). The total number of deaths by COVID-19 during a particular month (from February to December) was our dependent measure.

The results indicated a significant effect of %vac (b=-2.78, χ^2^(1)=34.75, p=3.75.10^−9^), a significant effect of #cases (b=0.90, χ^2^(1)=30.94, p=2.66.10^−8^), a significant effect of time2 (b=1.58, χ^2^(1)=9.51, p=0.002), and a significant effect of the interaction between %vac and #cases (b=-0.01, χ^2^(1)=34.46, p=4.36.10^−9^); the effect for the linear trend (time) wasn’t significant (b=0.54, χ^2^(1)=0.01, p=0.937). We retained the time covariate for its complementary effect (with respect to time2) on the quadratic polynomial trends in the following models. It should be noted, however, that excluding the time covariate leaves all other effects intact with respect to their directions, magnitudes and significance levels.

Note that the fixed effects for %vac and #cases make sound theoretical sense: the number of deaths decreases as the percentage of vaccinated people increases while the number of deaths increases with the increase of the total number of COVID-19 positive cases occurring during the previous month.

The variances/covariances of our random effects are given in table 1:

**Table 1.** Variances (main diagonal, bold) and covariances (below the main diagonal) between the 3 random effects specified for our baseline model. The error variance for the whole model was equal to 3411.08. AIC=3118.8.

|  | *Intercept* | *%vac* | *#cases* |
| --- | --- | --- | --- |
| *Intercept* | **2984** |  |  |
| *%vac* | -41.40 | **0.73** |  |
| *#cases* | 24.42 | -0.30 | **0.21** |

We investigated the covariance structure of the baseline model by means of both Likelihood Ratio (LR) Tests and Akaike Information Criteria (AIC). Results indicated that all random effects included in the baseline model were highly significant: all ps from LR Tests were <0.0003 and all ΔAICs were ≥13 in favor of the baseline model being compared to a trimmed version of it.

Next, we investigated the usefulness of including time and time2 as random factors in the model. The results were as follows:

- Including time as a random effect did not improve the overall model significantly (p=0.141, ΔAIC=1.1 in favor of the baseline model);
- Including time2 as a random effect did not improve the overall model significantly (p=0.111, ΔAIC=0.5 in favor of the baseline model);
- Including both time and time2 as random effects did not improve the overall model significantly (p=0.446, ΔAIC=9.1 in favor of the baseline model);
- Finally, we entered time and time2 as random effects but separately, i.e. they were allowed to covary between themselves but not with the random effects already specified within the baseline model, thereby exploring a partially diagonal, partially unstructured covariance matrix. The results indicated that this approach also appeared not to improve over the baseline model (p=0.97, ΔAIC=6 in favor of the baseline model).

We should also mention that several of the above approaches resulted in convergence difficulties indicating once more that random effects pertaining to time and time2 were indeed superfluous. Also, it is worth noting that all changes of the baseline model which we attempted didn’t change appreciably the fixed effects of time2, %vac, #cases and their interaction (i.e. their directions, magnitudes and significance levels remained virtually the same regardless of the random effects added to the baseline model) described above.

In summary, our baseline model seems to fit the data better than others and our main observation within its context is the significant effect of the percentage of vaccinated people on the total number of deaths recorded during the following months. The coefficient for %vac is -2.78 meaning that each percentage point increase in vaccination is associated with the decrease of more than two deaths per 1M in the general population on average across EU countries (holding all other variables and interactions in the model constant). We should note that the average number of deaths due to COVID-19 (per 1 M) across the 12 months during 2021 for all EU countries was equal to 119.87. Note that for most countries and for most months the vaccination rate increases with more than 1% per month and that the -2.78 effect is a cumulative one, i.e. it accumulates over all measured time points (i.e. months) so the overall effect of the vaccination campaign should be expected to be much higher as we try to illustrate below. We should also note that, consistent with all theoretical expectations, the number of COVID-19 positive cases recorded during a particular month correlates positively with the number of COVID related deaths for the following month. This observation should serve to increase the trustworthiness of the presented model (also the #cases effect serves as proper control with respect to the vaccination effect).

Having established some usefulness of our baseline model, we will proceed to examine some of its specifics. After calculating each country’s specific intercept and slopes^6^ (for %vac and #cases), we correlated the results and observed the following patterns:

- The slopes for all countries with respect to %vac were negative indicating that for each individual EU country the number of deaths due to COVID-19 was estimated to decrease with the increase of the relative number of vaccinated individuals. The slopes’ coefficients ranged between -5.36 (Hungary) and - 1.89 (Netherlands);
- The slopes for all countries with respect to #cases were positive indicating that death rates increased with the number of infectees. These coefficients ranged between 0.32 (Sweden) and 2.11 (Hungary);
- There was a significant correlation between the total number of vaccinated people up to and including December 2021 (vac_tot) and the intercepts calculated for each country: r=-0.57, p=0.003. This indicates that populations vaccinating at higher (and presumably faster) rates tend to experience less COVID-19 casualties overall. This observation corroborates the findings with respect to the %vac effect reported above;
- There was a significant positive correlation between vac_tot and the slopes for %vac (r=0.54, p=0.005). Since all slopes for %vac were negative, this means that countries with higher (faster) rates of vaccination tend to benefit less from the vaccines than countries with lower (slower) overall rates of vaccination. Various explanations for this observation are possible. For example, this may be an artifact from highly vaccinated populations showing lower slopes of %vac because as the rate of vaccination approaches 100%, its variance in time declines and hence its explanatory power decreases (in other words, once nearly 100% of a population is vaccinated, its rate of vaccination stops changing in time and hence COVID-19 deaths or lack thereof cannot be predicted on the basis of vaccination rate, i.e. a constant has no predictive power). Another possible reason for the above observation may be that populations with faster rates of vaccination are those which vaccinated many people in the early months of 2021 and due to vaccines’ proposed waning effectiveness, these populations were more exposed to different COVID-19 variants occurring in the later 2021 months, thereby exhibiting diminished estimates of the vaccines’ overall effectiveness. These and other possible explanations for the discussed phenomenon definitely deserve further investigation;
- Finally, we observed a significant negative correlation between vac_tot and the slopes for #cases (r=-0.57, p=0.003). This association is particularly interesting for it shows how the vaccination rate moderates the effect #cases exerts on our dependent measure (monthly mortality rates). Specifically, we see that more vaccinated populations exhibit diminished relationships between the number of newly diagnosed COVID-19 cases per month and the number of COVID-19 casualties for the next month in comparison to less vaccinated populations. Thus, the rate of vaccination appears not only to reduce casualties directly, but also indirectly, through breaking down the dependence of casualties on the number of infections. This is exactly what one would expect to see if vaccination reduces the risk of complications and death due to COVID-19. Please compare this effect to the negative covariance between the random slopes for #cases and %vac reported in Table 1 and to the significant interaction between the fixed effects of %vac and #cases described at the beginning of this section.

### 3.1. An illustrative case study

Let’s illustrate the effect of vaccination by a case study involving the most vaccinated and the least vaccinated countries in the EU. Fig. 1 illustrates the time trajectories for Bulgaria (the least vaccinates country in the EU with vaccination rate of 27.61% by December 31^st^, 2021) and Portugal (the most vaccinates country with 90.5% vaccination rate by the same date).

**Figure 1.**
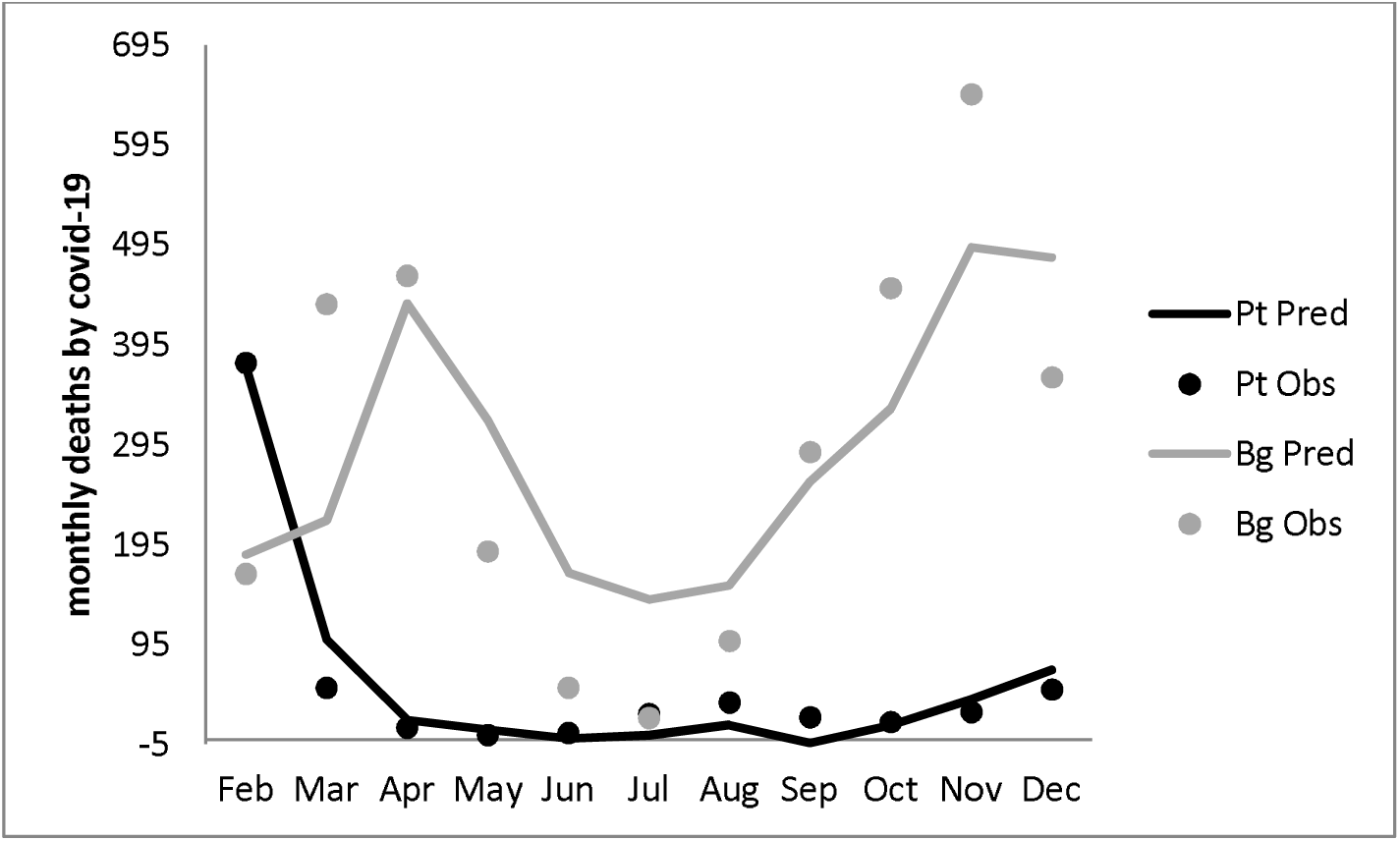
Time trajectories for Bulgaria (Bg) and Portugal (Pt) with respect to COVID-19 monthly mortality rates. The dense lines represent the values fitted by our baseline model; the dots represent the actual data points. The fitted trends (dense lines) were based on each country’s individual slopes and intercepts calculated by the baseline LMEM.

Fig. 1 illustrates the very different outcomes for the two countries. It should be noted that Bulgaria’s extraordinary death rate doesn’t appear to be due to more infected cases per month on average. On the contrary, for Portugal, the total number of infectees (calculated from January to November for this is the data used for predicting the dependent measure) is equal to 721 (per 10K) while for Bulgaria the same number equals 715. Obviously the two rates are almost identical (with Portugal actually showing more infected patients per capita than Bulgaria). At the same time, Bulgaria’s total mortality rate from February to December (data points included in our dependent measure) equals 3177 while Portugal’s number amounts to only 637 (per million). To illustrate this point further we present another figure (Fig. 2) which uses the same data with the sole exception that Bulgaria’s monthly vaccination rate is replaced by Portugal’s (all other data points for the two countries remains intact).

**Figure 2.**
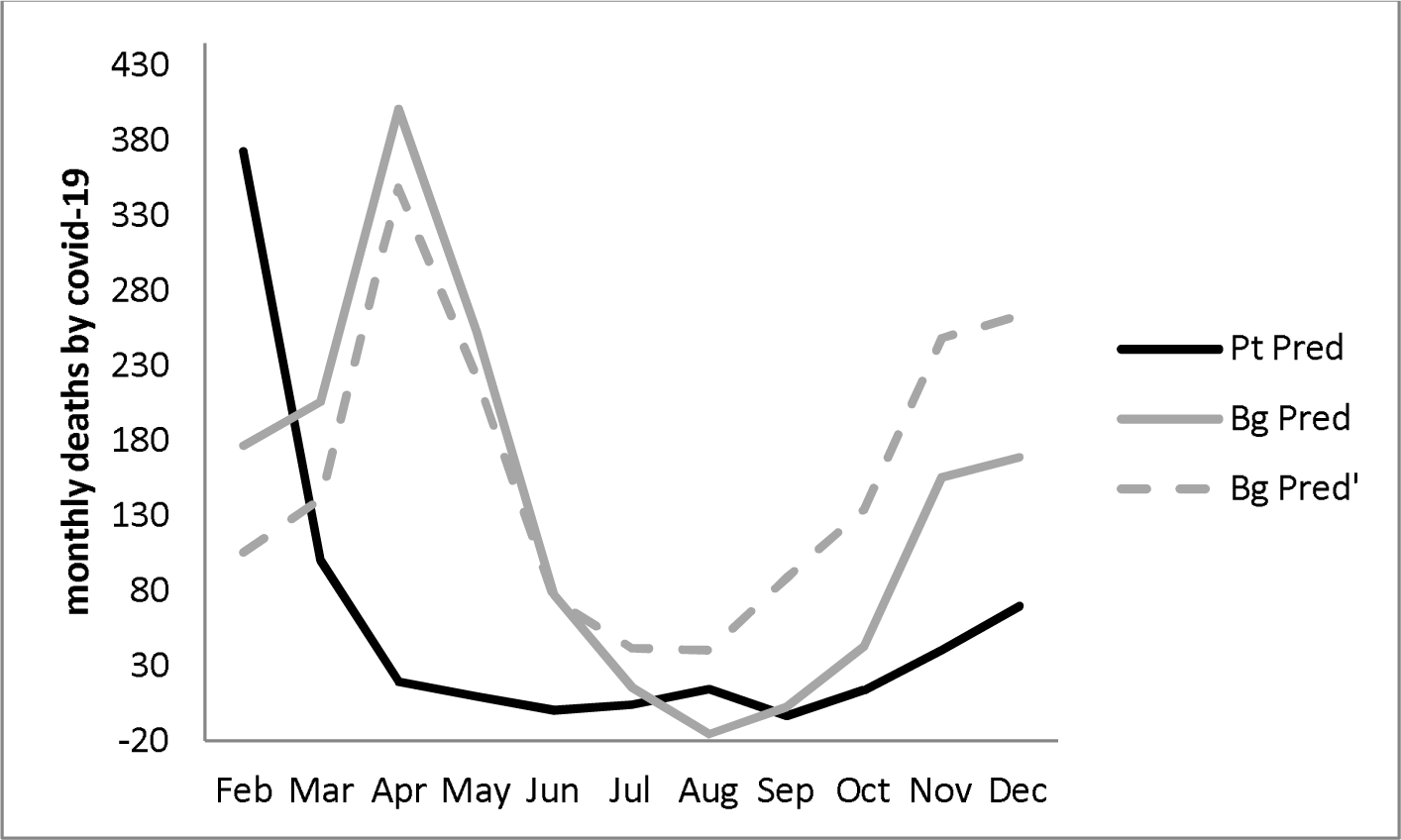
The predictions of our baseline model for the hypothetical case where Bulgaria’s vaccination rate is exactly as Portugal’s. #cases and all other variables/interactions are kept intact. The individual slopes and intercepts for both countries are also the ones obtained from the original analysis. The dashed line (Bg pred’) gives prediction for Bulgaria’s mortality rates based on Portugal’s vaccination dynamics and Portugal’s individual slope while all other data points are kept intact.

With our actual data, the baseline model predicts total mortality rates (February to December) of 3185 and 642 per million for Bulgaria and Portugal, respectively (compare with the actual values of 3177 and 637). Replacing Bulgaria’s vaccination values for the first 11 months of 2021 with Portugal’s however, causes our model to predict only 1485 COVID-19 casualties for Bulgaria for the same period. It may be argued that since we are using Portugal’s vaccination rates in this simulation, we should also adopt Portugal’s slope for %vac. The dashed line in Fig. 2 shows the results when we do so; this time the mortality rate predicted for Bulgaria is 1709, still much lower than both the actual number and the model prediction based on Bulgaria’s actual vaccination values. We see that %vac appears to have a pronounced cumulative effect with respect to mortality rates.

### 3.2. Additional controls

In order to explore the robustness of the above model we entered several fixed (i.e. not changing through time but only from country to country) covariates. More specifically, we entered the following as fixed control measures: population density, percentage of people over the age of 65, gross domestic product (gdp) per capita, cardio-vascular death rate, diabetes prevalence, hospital beds per 1000 people, and life expectancy. None of these additional control measures seemed to exert any significant influence on the dependent variable (all ps>0.40). Also, AIC seemed to favor our baseline model over the one including the control variables in question: ΔAIC=8.7 in favor of the baseline model; the LR Test also suggested that including the seven control measures doesn’t improve the baseline model as a whole significantly: χ^2^(7)=5.373, p=0.615. That is not to say that the fixed variables don’t influence the dependent one, what the analysis shows is that the additional controls don’t seem to exert significant influence over the dependent measure once %vac, #cases and their interaction *have been taken into account*. It may well be the case that, say, gdp (per capita) as an indicator for quality of life influences the number of infected cases (#cases) which in turn influence mortality rates. However, once #cases has been accounted for, gdp has little if anything to add in terms of additional (i.e. over and above #cases’) explanatory power.

More importantly, all of the effects which were significant within the baseline model retained both their significance and their directions of influence (i.e. signs) in the presence of the seven additional control variables. For example, %vac was estimated as b=-2.27, p=1.21.10^−^5 within the augmented model.

### 3.3. HDI as an Additional Control

Having discarded the augmented model in favor of the baseline, we proceeded to investigate the Human Development Index^7^ (HDI) as an additional control measure. HDI is defined as the geometric mean of Life Expectancy at Birth, Education Index, and Gross National Income per capita (very similar to gdp). As can be seen, two of the three variables comprising the HDI were present within the 7 controls introduced above. HDI is supposed to reflect an aggregate level of a society’s well-being as well as the functional adequacy of various institutions (education and healthcare systems, different economic organizations, etc.). With entering HDI to our predictors we hoped to provide a more general explanatory framework with respect to societies’ preparedness to cope with the COVID-19 crisis. Also, at a more technical level, including an integrated measure instead of its subparts was an attempt at reducing multicollinearity (e.g. Life Expectancy and gdp correlated 0.625, p=0.000 for 2019). So, our next model contained HDI as a fixed predictor alongside the predictors already present in the baseline model.

Results indicated that HDI failed to reach significance (b=-3.3, p=0.137) although it appeared more promising than all fixed covariates described above (entering HDI into the equation didn’t change the significance of any of the time-varying predictors already present in the baseline model). Here, however, an interesting observation occurred: HDI correlated significantly with the random slopes for #cases for different countries calculated within the baseline model, r=-0.86, p=2.47.10^−8^. In other words, for countries with higher HDI’s the relationship between infection and mortality rates was smaller than for countries with lower HDI’s. This makes intuitive sense: countries with more developed infrastructure probably have greater capacity to cope with the burden COVID-19 imposes on their healthcare systems than countries with less social and economic resources. As we already saw above, this is a classic symptom of an interaction. Hence, we retained the HDI variable in the current model and included an interaction term for HDI and #cases. The results confirmed our speculations and are summarized in Table 2.

**Table 2.** Fixed and random effects for the model including the HDI and its interaction with #cases. The first (left) half of the table shows the estimates of the fixed predictors and their interactions (signified by *), their standard errors and associated t-values and their significance levels (based on LR Tests). The second (right) half of the table shows the variance/covariance estimates of the random effects included in the model as well as the AIC value.

|  | *Estimate*<br>*(b):* | *Standard*<br>*Error:* | *t-value:* | *p:* |  | *Intercept* | *%vac* | *#cases* |
| --- | --- | --- | --- | --- | --- | --- | --- | --- |
| **Intercept** | 62.224 | 21.119 | 2.946 | 0.005 | **Intercept** | 1290 |  |  |
| **%vac** | -2.438 | 0.438 | -5.565 | $3.54 \cdot 10^{-7}$ | **%vac** | -24.744 | 0.599 | |
| **#cases** | 0.849 | 0.081 | 10.465 | $3.87 \cdot 10^{-12}$ | **#cases** | 8.302 | -0.140 | 0.057 |
| **time** | -3.383 | 6.619 | -0.511 | 0.619 |  |  |  |  |
| **time2** | 1.749 | 0.500 | 3.494 | 0.001 | **Residual** | 3391 |  |  |
| **HDI** | -9.895 | 2.040 | -4.850 | $5.37 \cdot 10^{-5}$ | **AIC** | 3100.8 | | |
| **%vac\*#cases** | -0.011 | 0.002 | -5.098 | $8.53 \cdot 10^{-7}$ | | | | |
| **HDI\*#cases** | -0.101 | 0.019 | -5.378 | $8.24 \cdot 10^{-6}$ | | | | |

We see that both HDI and its interaction with #cases are now highly significant while the effects inherited from the baseline model remain unchanged^8^. In other words, the interaction term between HDI and #cases appears to play a crucial role in the model as a whole (e.g. including this interaction term renders the main effect of HDI highly significant as opposed to the case when only the HDI effect was present in the model). It appears that indeed HDI moderates the relationship between #cases and mortality rates with countries exhibiting higher HDI’s showing lower dependence of mortality rates on infection rates than countries with lower HDIs, presumably because more developed countries have the resources to withstand the burden COVID-19 poses on their healthcare systems and other social infrastructures^9^.

We should point out that HDI correlates significantly (r=0.69, p=1.25.10^−4^) with the total number of vaccinated people up to and including December 2021 (vac_tot) which was discussed in (3). Several reasons might lay at the heart of this phenomenon including richer countries finding it easier to obtain enough vaccines during the early periods of the pandemic (this, however, is unlikely to be the case since the EU employed a common policy for ordering, buying and distributing the vaccines), better education increasing the probability of people taking the shot, and better functioning institutions finding better ways of convincing (or even coercing) citizens to take the shot among others. Whichever the case, it appears that HDI explains a large portion of the variance between countries with respect to vaccination rates (this might be the subject of a separate analysis). Our current data supports the above conclusion in the following way: as we saw in (3) vac_tot correlated significantly with all random effects (intercepts, slopes for %vac and slopes for #cases) within the baseline model. Here, however, it appears that once HDI and its interaction with #cases were accounted for, the correlations between vac_tot and the random effects for each country disappear – all ps>0.10. In other words, it appears that the correlations found in (3) were the result of a common cause (HDI) being excluded from the model. Once HDI was included and both its moderation effect on the relationship between #cases and mortality rates and its relationship to vaccination rates were accounted for, the interrelations between vaccination rates and the random effects tend to diminish appreciably. This observation suggests that more developed countries tend to vaccinate their populations at higher (faster) rates while also being capable of withstanding the burden COVID-19 places on different institutions to higher extents. These two tendencies are, in all likelihood, related for higher vaccination rates appear to reduce the number of hospitalizations, length of hospital stay and ICU admissions (e.g. Whittaker et al., 2021). Also, the somewhat puzzling negative correlation between vac_tot and the slopes for %vac reported in (3) disappears once HDI is included in the model.

Fig. 3 shows the predicted mortality trends for 10 EU countries based on our HDI-containing model.

**Figure 3.**
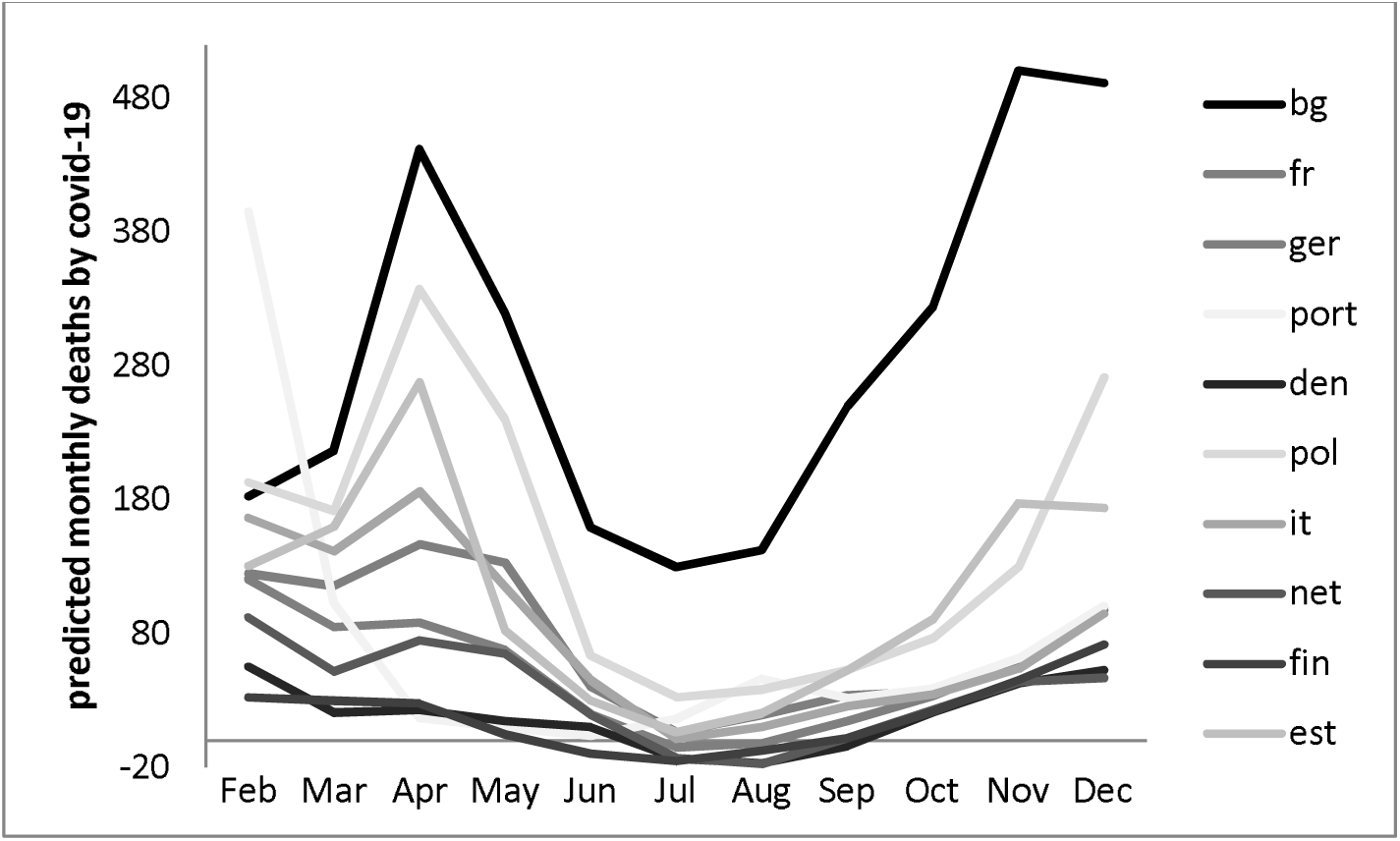
Predicted monthly deaths by COVID-19 for 10 EU countries on the basis of the model containing HDI and its interaction with #cases. The data is for Bulgaria, France, Germany, Portugal, Denmark, Poland, Italy, Netherlands, Finland and Estonia. The trajectories were calculated taking into account the individual intercepts and slopes (for #cases and %vac) for each different country.

Finally, we included the effect of the total number of deaths due to COVID for the previous month as a predictor to the mortality rates during the next month. This approach brings us as closer to a genuine longitudinal approach as possible in the current context and involves a new control variable which should be strongly related to the dependent measure (for the two measure the same thing with a time lag of one month).

The results indeed indicated that mortality rates for the previous month had significant predictive power: b=-0.27, p=1.1.10^−4^. The negative sign of the parameter estimate shows that mortality rates for the next month are negatively influenced by previous mortality rates. This may result from natural fluctuations in the virus spreading (including seasonal variations, acquired immunity, etc.), institutional reactions to heightened mortality rates (e.g. quarantine measures) or it can be an artifact from the so called negative suppression (e.g. Maasen & Bakker, 2001) which is occasionally encountered in regression analyses.

More importantly, the inclusion of previous months’ mortality rates didn’t change any of the effects discussed so far either in terms of their significance or with respect to their directions (e.g. for %vac we have b=-2.82, p=1.32.10^−7^), thus reinforcing the notion that the effects discussed so far are indeed robust.

## 4. Discussion

Several mixed-effects models indicated that the cumulative percentage of vaccinated citizens within a country is negatively related to mortality rates.

All models produced estimates for the linear effect of %vac ranging between -2 and -3 deaths per million per month on average. As we saw in (3.1) however, the overall effect of vaccination through time can be quite dramatic.

We should again emphasize that different results may be observed in the context of different time lags. It also remains unclear the extent to which our approach has been able to adequately account for the obvious nonlinearities in the time trends. Including time squared as a covariate as well as several interaction terms obviously produces nonlinear time trajectories but we should be cautious in our interpretation because of the obvious complexity of mortality rates’ dynamics.

It is also questionable whether our findings will extend to countries outside the EU.

It is clear that two models appear to provide good fits to our data – the baseline model and the model adding HDI as a fixed covariate alongside its interaction with the monthly number of newly diagnosed cases. Both models appear to fit the data well, both models include the same random parameters (random intercepts as well as random slopes for %vac and #cases included in an unstructured variance/covariance matrix). It is also the case that the two models make very similar predictions with respect to countries’ individual time trajectories (i.e. the two models provide very similar estimates for the random intercepts and slopes for the different countries). While it is true that HDI appears to be involved in two significant effects, we should be cautious about possible overfitting. Currently, we would like to refrain from definite conclusions in that respect until more data is collected, analyzed and cross-validated.

It is also possible that our analyses underestimate the effect of the vaccines due to another reason. Although we included #cases in all analyses and therefore partialled out its effect from the vaccines’ effects, we didn’t control for the fact that vaccination, in all likelihood, reduces #cases across certain time lags. Our analyses focused on predicting mortality rates one month after particular %vac and #cases had occurred. In other words, #cases and %vac as predictors are measured for the same time period. It may well be the case, however, that vaccines have indirect effects on mortality rates through their influence on the infection rates. For example, it may be the case that %vac at time t influences #cases at time t+1, which in turn influences mortality rates at time t+2; since mortality rates at time t+2 are obviously influenced by %vac at time t+1, it may be suspected that the total effect of %vac (i.e. the direct effect of %vac at time t+1 on mortality rates at time t+2 plus the indirect effect of %vac at time t on #cases at time t+1 which influences mortality rates at time t+2) be higher than the effects reported above. A preliminary analysis attempting to address this issue is presented in the next section.

## 5. Growth Curves Analysis

We used an Individual Growth Curves Analysis (IGC) in order to test for potential indirect effects of vaccination rates on mortality rates through infection rates. Also, this approach is similar to LMEM (e.g. Bollen, 2005).

IGC typically models individual intercepts and time related slopes as latent variables with fixed loadings on the actual measurements; time-varying covariates are specified at the next levels of the analysis with the overall structure resembling classical LMEM.

The relatively small sample size with respect to measurement units (25 countries) precluded us from making use of our entire dataset. Hence, we decided to choose 4 waves of measurements to include into the analysis. Concretely, we chose measurements ranging from August to October with respect to %vac and #cases and from September to November with respect to monthly mortality rates (thus we employed the same one-month-lag procedure present in our previous analyses). The choice of this particular time period reflected the following considerations:

- The variance for %vac was highest for September and we decided to include this point in the analysis because, all else being equal, variables with larger variances have (potentially) higher explanatory power. In hindsight, this particular reasoning may have resulted in some problems as we shall see shortly;
- We wanted to concentrate on months towards the end of 2021 whereby most countries had achieved vaccination rates capable of making a measurable impact on the substantial infection rates present during the period;
- We decided to exclude December since during this month the OMICRON variant became dominant in several (former) EU countries. Since OMICRON appears to exhibit marked deviations from previous variants (e.g. larger capacity to bypass the vaccines, milder course of the disease, reduced incubation period), we didn’t include December in the analysis in order to avoid potential sources of confounding.

Prior to the analysis we specified the causal structure shown in Fig. 4. As will be shown shortly, this basic structure underwent some changes necessary for the model to fit the data adequately.

**Figure 4.**
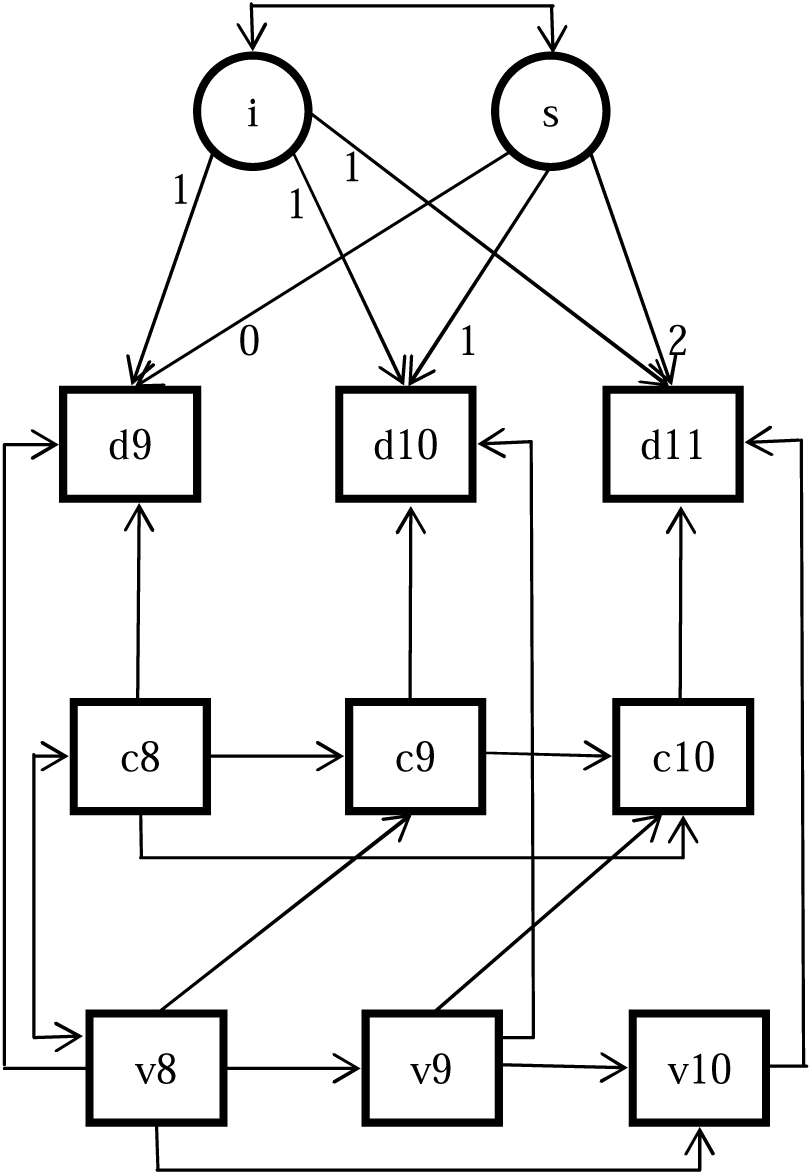
The intended causal structure of our IGC. “v”, “c” and “d” denote %vac, # cases and monthly death rates respectively. The numbers (8 to 11) denote the data for particular months (e.g. 8=August, etc.). Observed variables are represented as rectangles while the latent intercept and slope (i and s at the top) are encircled. Fixed parameters are denoted by numbers representing the values parameters were fixed on. Double arrows denote (unanalyzed) variances; single arrows denote path coefficients. The two indirect effects which we were primarily interested in were the ones from v8 to d11 through c9 (i.e. v8->c9->d10) and from v9 to d11 through c10 (i.e. v9->c10->d11).

We began by testing the intercept/slope structure of the model with respect to mortality rates (the top two levels of Fig. 4). The model was estimated via the standard Maximum Likelihood algorithm and results indicated a very good fit – χ^2^(1)=1.05, p=0.306, CFI=0.999, RMSEA=0.044 (95% CI=[0.000 – 0.534]), SRMR=0.052. We see that a random intercept/slope with a linear time trend only fits the three monthly mortality measurement waves very well. Since we included only three time points in this analysis, it is hardly surprising that we didn’t have to include the polynomial time trend necessary in the context of the LMEMs described above.

After having established the viability of the measurement portion of the model, we proceeded to including the lower levels time changing covariates.

Fig. 4 shows the structure which was initially proposed. Note that save for the latent random intercept/slope structure (top), the rest of the variables were involved in a pattern resembling typical longitudinal path analyses (e.g. Kline, 2010). As the caption below Fig. 4 explains there were two indirect effects which we intended to test – from %vac up to and including August (v8) to mortality rates for October (d10) through infection rates (September) and from %vac September (v9) to mortality rates November (d11) through infection rates October.

Our initial results suggested a medium fit to the data and inspection of the modification indices clearly showed that there were a few sources of noticeable strain on the model. After adding three coefficients suggested by the largest fit indices (Fig. 5) we obtained an excellent fit to the data.

**Figure 5.**
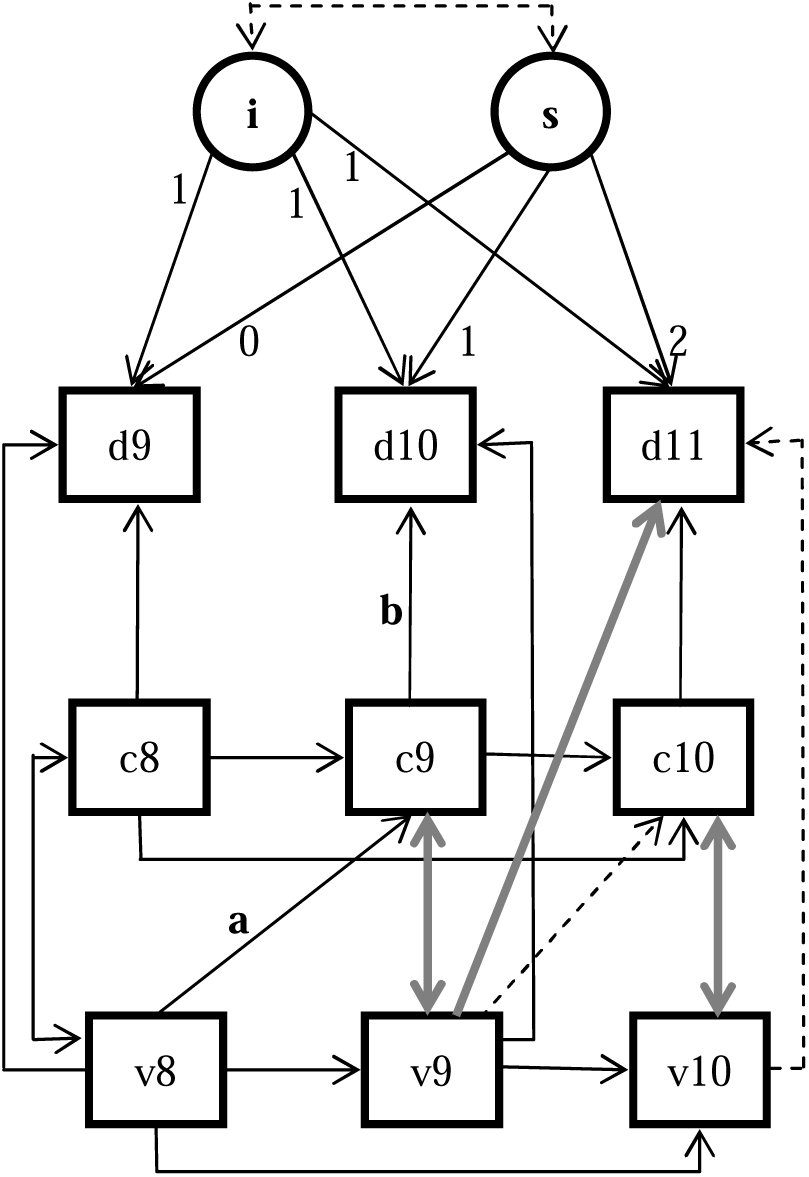
Graphical representation of some results obtained from our IGC analysis. The grey lines represent error covariances and a path coefficient suggested by the modification indices; all of these were significant, ps<0.05. The dense lines (grey and black) represent significant (all ps<0.05) path coefficients and (error) covariances. The dashed lines represent originally specified relationships which didn’t reach significance within the final model depicted in the figure. All significant path coefficients were in the expected directions. The “a” and “b” signify the significant indirect effect of cumulative vaccinations up to and including August on monthly mortality rates for October through November infection rates. See the text for more details

Specifically, χ^2^(16)=15.303, p=0.503, CFI=1.000, RMSEA=0.000 (95% CI=[0.000 – 0.178]), SRMR=0.022.

Several things are worth noting with respect to the final model (Fig. 5):

- All significant path coefficients were in the expected directions, i.e. all significant coefficients from %vac to mortality rates were negative (ranging from -3.6 for the v8->d9 coefficient to -15.4 for the v9->d11 coefficient^10^); also, all significant paths from #cases to mortality rates were positive (ranging from 0.35 for c10->d11 to 1.07 for c9->d10); finally, there was a significant path from %vac to #cases (v8->c9);
- We didn’t observe a significant indirect effect of v9 on d11 through c10. Fig. 5 clearly shows the reason why: on one hand the effect of v9 on c10 doesn’t reach significance, on the other v9 appears to have a significant *direct* effect on d11 as suggested by the modification indices (the direct effect, of course, tends to diminish the importance of the indirect ones all else being equal);
- Two out of the three one-month lagged effects of %vac on mortality rates are significant with the third effect (i.e. v10->d11) failing reach significance presumably because v9 appears to have a strong effect on d11 (i.e. v9 appears to influence d11 via a two-months lag and v10 obviously has little to add in terms of explanatory power once v9 is accounted for; as we already mentioned v9 is the %vac measurement with the highest variance for 2021 and in that sense the above observations are not entirely surprising). Also, it appears that all one-month lagged effects of #cases on mortality rates are significant and in the predicted direction with no additional parameters (e.g. two-months lags) needed in order to account for the data at hand. These observations tend to corroborate the findings discussed in the context of the LMEM approach;
- Our IGC model finds significant paths from v8->c9 and from c9->d10; hence, it should come as no surprise that the indirect effect of v8 on d10 through c9 is significant (b=-1.54, p=0.011)^11^. Since v8’s direct effect on d9 was equal to - 3.6 (p=0.000), it seems that its indirect effect on d10 through c9 was almost half as large as its direct effect.

In summary, the results from the IGC analysis seem to corroborate our previous findings. In general, the LMEM’s estimates of the effects in question should be regarded as more reliable than the ones obtained from this preliminary IGC model since the linear-mixed procedure makes use of our entire dataset (as opposed to the small subsample included in the current analysis) and deals with more data points per measurement unit (country) thereby making use of much more reliable information with respect to individual time trends. That being said, IGC suggests that both concerns of our LMEM underestimating the actual effects of the vaccines may have indeed been reasonable. First, it appears that the one month lag may not always be the optimal time period for assessing the vaccines’ effectiveness for as we just saw, it appears that v9 has a large significant effect on d11, i.e. an effect being carried over two months instead of one (b=-15.4, p=0.005). Second, it does appear that vaccines have had discernible indirect effects for certain time periods on mortality rates through infection rates: one of the indirect effects tested within the IGC model proved significant and in the predicted direction.

Although the above results should be regarded as tentative due to the small sample size and restricted time period the analysis is applied on, it deffinately appears that the effects in question should be investigated further. In other words, it may well be the case that the vaccines’ effectiveness with respect to preventing fatalities due to COVID-19 reported in (3) was actually underestimated.

## 6. Conclusion

It definitely appears that vaccination against COVID-19 reduces mortality rates in the EU.

Our LMEM models suggested some interesting interaction effects. For example, it appears that HDI appears to moderate the relationship between infection and mortality rates, presumably at least partly due to more developed countries showing higher vaccination rates which in turn, in all likelihood, reduce the number of COVID-19 related complications, hospitalization rates and ICU admissions.

More studies should be performed on different and preferably larger datasets (while still ensuring the comparability of different populations’ estimates for vaccination, infection and mortality rates) in order to validate the above observations.

An Individual Growth Curve Analysis suggested that both indirect effects of vaccination rates on mortality rates through infection rates and (partially) misspecified time lags may have led to underestimation of the vaccines’ effectiveness. More studies involving larger datasets should address these issues further.

Currently it appears that the cumulative effect of vaccination in reducing COVID-19 mortality within the EU is substantial.

## Data Availability

All data produced in the present study are available upon reasonable request to the authors

## Conflicts of Interest

This work has received no financing from any private, corporate or governmental source. The author is not affiliated with any private, corporate or governmental entity except for the educational institution mentioned at the beginning.

## Footnotes

1 See “Report of the WHO-China Joint Mission on Coronavirus Disease 2019 (COVID-19): https://www.who.int/publications/i/item/report-of-the-who-china-joint-mission-on-coronavirus-disease-2019-(covid-19)

2 Note that the decision to use a monthly lag in this study was based on theoretical considerations and prior studies only, i.e. we made no attempt to find a lag which maximized the investigated relationship. In that sense, it may be expected that our estimates may be somewhat conservative since, in all likelihood, there are different lags which show more pronounced effects than the ones reported below. In that sense, it is probably a good idea to conduct an exploratory study which explicitly tries to find the time period which best exemplifies the relationship between vaccination and mortality rates and subsequently to validate the observed results across an independent sample. Since currently we focus only on EU countries and since the vaccination program only really took off at the beginning of 2021, we regard the above proposition as a future research goal.

3 https://ourworldindata.org/covid-vaccinations

4 Note that the time covariates are supposed to reflect seasonal trends and thus the time codes designate months of the year rather than simply increasing time periods. In that sense, extrapolations beyond the current dataset should involve repeating the time codes for the upcoming months (e.g. January 2022 should be coded as 0 rather than as 11).

5 More concretely, the means for the entire dataset (rather than for individual countries) were subtracted from a variable involved in an interaction term prior to analysing the data.

6 In other words we calculated three new variables – one containing the specific intercept for each country, another containing the specific slope for each country with respect to #cases, and yet another containing the specific slope for each country with respect to %vac. These coefficients have averages equal to the fixed effects reported above. Their variances however are not equal to the variances of the random effects observed in Table Indeed, the variances of the actual estimates are smaller, for these estimates exhibit “shrinkage”, i.e. they are closer to their respective means than suggested by the variance estimates (Table 1) due to the hierarchical nature of the linear mixed model (e.g. Fitzmaurice et al., 2011). This feature automatically corrects for certain possible overfitting effects, including (hopefully) the ones to be described.

7 See http://hdr.undp.org/en/content/human-development-index-hdi. Here we used the data available for 2019. Also, we transformed the raw data (varying between 0 and 1) into a scale varying between 0 and 100 in order to match the measure’s variance to the variance of our other variables; finally we centered the variable (i.e. subtracted the overall mean from each observation) before entering it to the analysis.

8 We should also emphasize that this model also suggests that the slopes for all EU countries with respect to %vac are negative and that all slopes of #cases are positive, i.e. the general pattern observed within the baseline model remains unchanged.

9 The possibility that the Education part of the HDI is related to people’s willingness to follow different safety protocols (e.g. wearing masks, keeping social distance, etc.) should also be considered in future studies.

10 This effect diminishes to the somewhat more reasonable -8.3 (p=0.000) estimate if the insignificant path v10->d11 (Fig. 5) is discarded without any noticeable deterioration of the model fit indices.

11 The indirect effect is found by multiplying the “a” and “b” paths shown in Fig 5 (a=-1.44, b=1.07, a.b=-1.54). In order to verify the indirect effect’s significance we also calculated its standard error by a standard (naïve) nonparametric bootstrapping procedure (Efron & Tibshirani, 1994); based on the results, its significance was equal to 0.017. The same procedure also attested to the good fit of our entire model – p=0.925 by the Bollen-Stein bootstrapping procedure (Bollen & Stein, 1992).

## Notes

### Competing Interest Statement

The authors have declared no competing interest.

### Funding Statement

This study did not receive any funding.

## References

Amit, S., Regev-Yochay, G., Afek, A., Kreiss, Y., Leshem, E. (2021). Early Rate Reductions of SARS-CoV-2 Infection and COVID-19 in BNT162b2 Vaccine Recipients. The Lancet, 397(10277), 875 – 877.

Bates, D., Maechler, M., Bolker, B., Walker, S. (2015). Fitting Linear Mixed-Effects Models Using lme4. Journal of Statistical Software, 67(1), 1 – 48.

Bollen, K., Stein, R. (1992). Bootstrapping Goodness-of-Fit Measures in Structural Equation Models. Sociological Methods & Research, 21(2), 205 –229.

Bollen, K. (2005). Latent Curve Models: A Structural Equation Perspective. Wiley-Interscience, 1st Ed.

Efron, B., Tibshirani, R. (1994). An Introduction to the Bootstrap. Chapman and Hall/CRC; 1st Ed.

Fitzmaurice, G., Laird, N., Ware, J. (2011). Applied Longitudinal Analysis. Wiley, 2nd Ed.

Kline, R. (2010). Principles and Practice of Structural Equation Modelling. The Guilford press, 3rd Ed.

Levin, E., Lustig, Y., Cohen, C., Fluss, R., Indenbaum, V., Amit, S., Doolman, R., et al. (2021). Waning Immune Humoral Response to BNT162b2 COVID-19 Vaccine over 6 Months. The New England Journal of Medicine, 385:e84.

Maassen, G., Bakker, A. (2001). Suppressor variables in Path Models: Definitions and Interpretations. Sociological Methods &Research, 30, 2001.

Hyams, C., Marlow, R., Maseko, Z., King, J., Ward, L., Fox, K., et al. (2021). Effectiveness of BNT162b2 and ChAdOx1 nCoV-19 COVID-19 Vaccination at Preventing Hospitalizations in People Aged at least 80 Years: A Test-Negative, Case-Control Study. Lancet Infectious Diseases, 21(11), 1539 – 1548.

Pouwels, K., Pritchard, E., Matthews, P., Stoesser, N., Eyre, D, et al. (2021). Impact on Delta on Viral Burden and Vaccine Effectiveness against New SARS-CoV-2 Infections in the UK. Nature Medicine, 27, 2127 – 2135.

Pilishvili, T., Gierke, R., Fleming-Dutra, K., Farrar, J., Mohr, N., Talan, D. (2021). Effectiveness of mRNA COVID-19 Vaccine among U.S. Health Care Personnel. The New England Journal of Medicine, 385:e90.

Public Health England, COVID-19 Vaccine Surveillance Report Week 38, 2021.

Rosseel, Y. (2012). lavaan: An R Package for Structural Equation Modeling. Journal of Statistical Software, 48(2), 1–36.

Singer, J., Willett, M. (2001). Applied Longitudinal Data Analysis: Modeling Change and Event Occurrence. Oxford University Press, 1st Ed.

Whittaker, R., Kristofferson, A., Salamanca, B., Golestani, E., Kvale, R. et al. (2021). Patient Trajectories among Hospitalized COVID-19 Patients Vaccinated with an mRNA Vaccine in Norway: A Register Based Cohort Study. medRxiv.

